# ED-Triage-Agent: A Framework for Human-AI Collaborative Emergency Triage

**DOI:** 10.64898/2026.02.17.26346501

**Authors:** Karthick Sharma, Harikrishnan Sivadas, Sandeep Reddy

## Abstract

Emergency Department triage is a critical decision-making process in which clinicians must rapidly assess patient acuity under high cognitive load and time pressure. We present ED-Triage-Agent (`ETA`), a multi-agent AI framework designed to augment clinical decision-making in Emergency Severity Index (ESI) classification through human-AI collaboration. The system operates in two phases: (1) autonomous patient intake via a conversational agent that collects structured symptom histories and (2) collaborative acuity assessment in which specialized agents prioritize patients for vital sign collection and generate ESI classifications with explicit clinical reasoning. Unlike monolithic AI prediction systems, `ETA` mirrors clinical workflow by supporting decisions at each triage stage while preserving clinician autonomy. We describe the system architecture, agent design principles, and a preliminary evaluation methodology using the ESI Implementation Handbook case studies (60 standardized cases). This work proposes a model for deploying multi-agent AI systems in time-critical clinical environments where explainability and human oversight are essential. Code and the evaluation framework are available at https://github.com/Karthick47v2/ED-Triage-Agent.

## 1 Introduction

Emergency department (ED) triage, the rapid assessment and prioritization of patients by acuity, represents one of the most consequential yet resource-constrained decision points in acute care. Effective triage determines which patients receive immediate attention and which can safely wait, but it occurs precisely when cognitive resources are most limited: during high patient volumes, staff shortages, and time pressure. In the United States alone, annual ED visits exceed 143 million [1], with global overcrowding contributing to preventable adverse events and increased mortality [2, 3].

The Emergency Severity Index (ESI), the dominant triage framework in North America, categorizes patients into five acuity levels based on clinical presentation and anticipated resource needs [4]. Despite widespread adoption, ESI based triage faces three fundamental challenges. First, single-clinician assessment creates bottlenecks and introduces substantial inter-rater variability, with clinicians achieving correct classification in only 59% of cases [5]. Systematic biases emerge across experience levels. Experienced clinicians undertriage (risking delayed critical care), while novice staff overtriage (misallocating resources) [6]. Second, triage operates as a single-point assessment that cannot adapt as patient conditions evolve during waiting periods, leaving high-acuity cases like myocardial infarction or sepsis at risk when presenting with initially stable vital signs. Third, the brief registration data available before vital sign collection provides limited information to guide prioritization, potentially leaving critical patients waiting in crowded queues while lower-acuity cases are assessed first. During high-volume periods, this cognitive burden becomes untenable. Additional strain arises from patients seeking routine care due to limited primary care access [7], as well as unpredictable surges from natural disasters or pandemics [3].

Early computational approaches attempted to address these limitations through rule-based expert systems and traditional machine learning models trained on historical triage data [8–10]. While these systems improved consistency, they proved brittle when handling edge cases, unable to process unstructured clinical narratives, and difficult to integrate into clinical workflows. Current digital triage tools remain largely administrative, consisting of electronic intake forms and basic alert systems, rather than providing intelligent cognitive support for clinical decision making.

Recent advances in large language models (LLMs) have demonstrated potential for triage decision support, with systems approaching experienced clinician performance on standardized vignettes [7, 11, 12]. However, despite these advances, clinical adoption remains limited. Existing AI triage systems exhibit critical limitations that hinder real-world deployment: most operate as end-to-end predictors that attempt to replace rather than augment clinician expertise, offering limited transparency into their reasoning and consequently undermining clinical trust [13].

More broadly, the gap between AI’s demonstrated potential and its actual clinical integration reflects challenges spanning data quality, workflow alignment, and accountability within safety-critical settings [14]. Critically, prevailing approaches assume the availability of complete clinical data such as vital signs before generating predictions, failing to accommodate triage’s inherently sequential, information-gathering nature in which prioritization decisions must often precede comprehensive assessment.

Multi-agent architectures offer a promising alternative paradigm. Unlike monolithic AI systems, multi-agent frameworks decompose complex workflows into specialized, collaborative components that mirror clinical team function. This enables transparent reasoning through explicit agent deliberation, preserves human oversight at critical decision points, and naturally accommodates distributed, sequential decision making. While recent work has explored multi-agent systems for medical tasks [15], their application to time-critical triage under partial information remains unexplored.

This paper introduces ED-Triage-Agent (`ETA`), a multi-agent framework that reconceptualizes AI’s role in emergency triage from replacement to augmentation. `ETA` operates in two distinct phases that mirror clinical workflow (Figure 1). Phase 1 (Pre-Triage Assessment) begins at patient arrival: an Intake Interview Agent conducts adaptive conversational symptom collection at self-service kiosks, implementing urgency-based question truncation to immediately flag high-acuity presentations. A Priority Assessment Agent then analyzes symptom data alone to generate provisional ESI estimates, enabling clinicians to identify which waiting patients require immediate vital sign assessment regardless of arrival order. Phase 2 (Post-Vital Classification) activates after clinicians collect vital signs and physical findings: specialized reasoning agents integrate the complete clinical picture to produce final ESI recommendations with explicit criterion-linked rationale.

**Figure 1:**
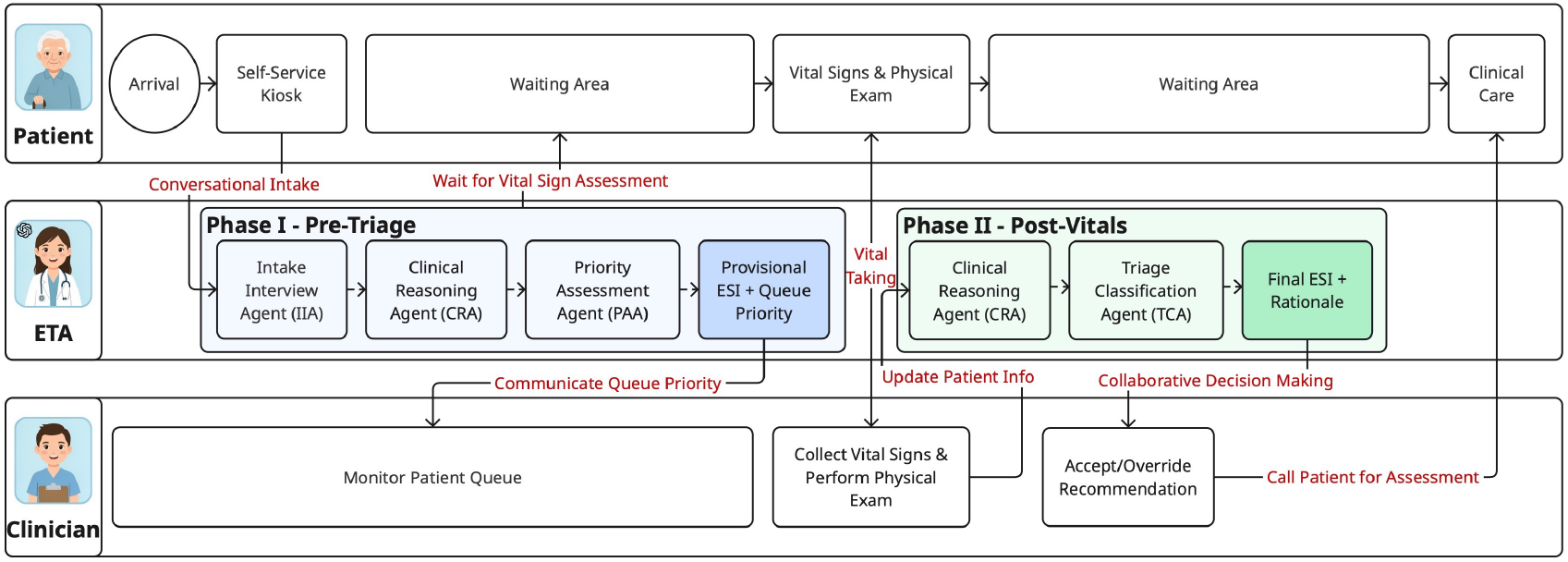
System overview of ED-Triage-Agent (`ETA`). Phase 1 supports early queue prioritization using symptoms alone, while Phase 2 generates final ESI recommendations after vital signs and clinical findings are available.

Our design contributions include: (i) a two-phase system architecture that mirrors actual ED triage stages, generating actionable recommendations at both pre-vital (queue prioritization) and post-vital (final classification) decision points; (ii) explicit preservation of clinician autonomy through interpretable, ESI criterion-linked reasoning with urgency-adaptive interview strategies that balance data collection against rapid risk identification; (iii) a complete end-to-end system specification including agent orchestration, knowledge grounding via RAG, safety mechanisms, and audit logging, providing a blueprint for clinical deployment; and (iv) a simulation-based benchmarking protocol using 60 ESI Implementation Handbook v4 standardized cases to assess system performance. We emphasize that `ETA` targets acuity assessment to support prioritization; we do not claim to replace clinical judgment for disposition or treatment planning, which remain exclusively within nursing and physician purview.

## 2 Background & Related Work

### 2.1 Emergency Severity Index (ESI) Triage

The Emergency Severity Index (ESI) is the dominant triage framework in North American emergency departments, categorizing patients into five acuity levels through a structured decision algorithm [4] (Table 1). ESI classification integrates two dimensions: clinical stability (risk of life threatening deterioration) and anticipated resource intensity (predicted number of diagnostic/therapeutic interventions).

**Table 1:** Emergency Severity Index (ESI) classification criteria.

| ESI Level | Category | Clinical Criteria | Resource Needs |
| --- | --- | --- | --- |
| 1 | Resuscitation | Requires immediate life-saving intervention | Any |
| 2 | Emergent | High-risk situation, confusion/lethargy/disorientation, or severe pain/distress | Any |
| 3 | Urgent | Stable, moderate symptoms | Many resources ($\geq 2$) |
| 4 | Less Urgent | Stable, mild symptoms | One resource |
| 5 | Non-Urgent | Stable, minimal symptoms | No resources |

The ESI algorithm proceeds sequentially. Clinicians first assess for immediate life threatening conditions (ESI-1), then evaluate for high-risk presentations or severe distress (ESI-2), and finally predict resource requirements for stable patients (ESI 3-5) based on expected resource consumption. This decision process requires synthesizing chief complaint, vital signs, pain assessment, mental status, and clinical judgment typically within 2-5 minutes per patient [3].

A critical gap emerges during overcrowding, clinicians must prioritize which waiting patients to assess first based solely on brief registration data (chief complaint, demographics) before vital signs are collected. Current ESI guidelines provide no systematic mechanism for this pre-vital prioritization, creating risk that high-acuity patients remain undetected while appearing externally stable.

### 2.2 AI Approaches to Triage Support

Computational triage support has evolved from rule-based expert systems [8] through classical machine learning approaches (logistic regression, random forests, gradient boosting) achieving 70-85% ESI classification accuracy [3, 9, 10, 16, 17]. However, these methods require extensive feature engineering, cannot process unstructured clinical narratives, and provide limited interpretability.

Deep learning approaches have demonstrated substantial improvements over classical machine learning for triage prediction. Convolutional neural networks (CNNs) and recurrent architectures such as bidirectional Long Short-Term Memory (BiLSTM) networks capture sequential dependencies and contextual patterns in clinical narratives, achieving higher discriminative performance for critical outcome prediction [9, 10]. While deep learning reduces reliance on manual feature engineering and can process raw clinical text, these methods remain constrained by requirements for complete structured inputs, limited interpretability that creates an “explainability crisis” hindering clinician trust, and susceptibility to dataset biases that may perpetuate disparities in risk assessment [3, 17].

Recent large language models (LLMs) demonstrate promising triage capabilities through natural language understanding and clinical reasoning. GPT-4 approaches resident physician-level performance on triage vignettes [11], while chain-of-thought prompting, few-shot learning and retrieval augmented generation improve reliability [7,1218]. Despite these advances, existing LLM based triage systems share critical limitations, they operate as end-to-end predictors requiring complete clinical data (including vital signs) before classification, failing to model triage’s sequential information-gathering workflow where prioritization must occur under partial information.

### 2.3 Multi-agent AI Systems

Multi-agent systems decompose complex tasks into specialized, collaborative components [19]. Unlike monolithic models, multi-agent architectures enable separation of concerns, specialized agent optimization for distinct subtasks, modular extensibility through agent addition, and concurrent execution for reduced latency [20]. Recent frameworks (AutoGen [21], LangGraph [22]) provide infrastructure for LLM-based agent orchestration with tool use, memory, and inter-agent communication.

Healthcare applications have explored multi-agent collaboration for clinical reasoning. MedAgents demonstrated collaborative diagnosis through multi-specialty agent discussion [15]. Two recent systems specifically address ED triage: TRIAGEAGENT employs confidence-weighted agent debates with retrieval augmented generation [23], while HEAL implements modular agents for information collection and continuous monitoring [24]. However, both systems assume complete clinical data availability before operation, lack pre-vital prioritization capabilities, and provide limited interfaces for clinician oversight, failing to address triage’s sequential, human-collaborative nature.

However, existing multi-agent triage systems exhibit limitations constraining clinical deployment. Both TRIAGEAGENT and HEAL assume complete clinical information availability before classification, failing to model sequential information gathering. Neither addresses pre-vital prioritization, determining which waiting patients need assessment first when queues accumulate. These systems lack explicit clinician integration, operating as autonomous collectives without clear interfaces for human oversight or override authority, raising concerns about clinical accountability. Additionally, multi-round deliberation architectures incur substantial computational costs through numerous API calls, increasing latency and operational expenses for time-critical applications.

No existing system supports triage decisions under partial information, enables pre-vital queue prioritization, or explicitly integrates clinician oversight at multiple workflow stages. `ETA` addresses these limitations through a two-phase multi-agent architecture aligned with clinical workflow.

### 2.4 Human-AI Collaboration Considerations

Effective clinical AI deployment depends on well designed human–AI collaboration. Transparency, calibrated trust, and preserved human agency are critical for adoption in safety critical settings [25]. Even highly accurate black-box models face resistance, as clinicians require interpretable reasoning that can be validated against domain expertise [13].

An augmentation paradigm, where AI supports rather than replaces clinicians, yields superior outcomes by offloading routine subtasks while retaining human decision authority [26]. Clinicians contribute contextual judgment, experiential intuition, and sensitivity to atypical cases that challenge algorithmic predictions. Effective systems minimize extraneous cognitive load while preserving germane load that supports skill development [27].

Automation bias poses a major risk, systems should promote critical engagement through explicit rationales, uncertainty communication, and deliberate friction before recommendation acceptance [28].

## 3 Methodology

`ETA`is guided by three core principles distinguishing it from prior AI triage systems:

### Augmentation over replacement

The system supports clinician decision making through structured information synthesis and calibrated risk estimates rather than autonomous decisions. Clinicians retain full authority over final ESI assignments.

### Workflow alignment

Rather than requiring complete clinical data before operation, `ETA` mirrors triage’s sequential information gathering nature, providing decision support at each stage including the critical pre-vital period when clinicians must prioritize assessment order with limited information.

### Transparent reasoning

All recommendations include explicit rationale linked to ESI handbook criteria, enabling clinicians to verify system logic against professional knowledge and identify cases where reasoning may be incomplete.

### 3.1 Multi-Agent Architecture Overview

`ETA` employs an orchestrator based multi-agent architecture where a central Orchestrator Agent (OA) manages workflow state transitions and routes information between four specialized agents (Figure 2). This deterministic orchestration pattern offers advantages over debate based architectures such as predictable workflow execution aligned with clinical stages, reduced latency through sequential processing rather than iterative deliberation, and clear accountability with traceable agent contributions.

**Figure 2:**
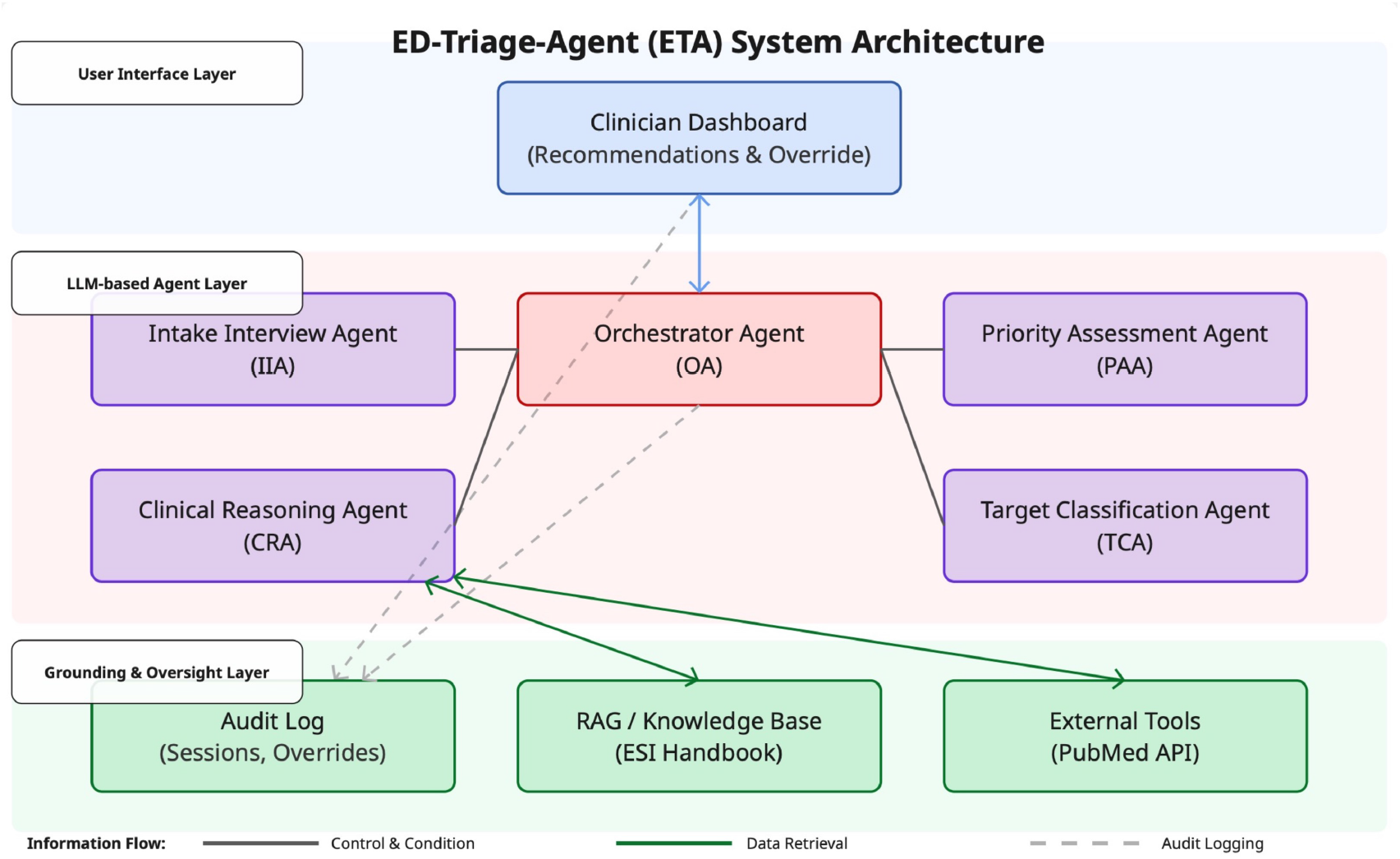
Multi-agent architecture of ED-Triage-Agent (`ETA`), showing the orchestrator, specialized agents, grounding resources (RAG / knowledge base), external tools, and audit logging with clinician dashboard oversight.

The system is implemented using LangGraph [22], enabling stateful workflow management with conditional routing. When the Intake Interview Agent detects high-acuity indicators, the orchestrator immediately flags patients for clinician assessment, bypassing standard workflow to prioritize safety.

`ETA` employs heterogeneous model selection based on task complexity and operational constraints. We selected the GPT-4.1 family as our foundation due to its superior instruction-following capabilities, enhanced performance on agentic tasks requiring multi-step reasoning. Within this family, we deploy GPT-4.1-mini for agents requiring clinical reasoning (CRA, PAA, TCA, OA), as it provides strong analytical capabilities for medical knowledge synthesis, risk assessment, and structured decision-making while maintaining reasonable latency and cost profiles. For the patient-facing conversational agent (IIA), we utilize GPT-4.1-nano, the most lightweight variant optimized for low-latency responses essential for natural patient interaction. This agent primarily performs information extraction from conversational exchanges rather than complex clinical inference, making the reduced capability acceptable while significantly decreasing response time and API costs. All agents access OpenAI models via Azure OpenAI API with temperature 0 to ensure deterministic, reproducible outputs appropriate for clinical decision support.

### 3.2 Two-Phase Workflow

A fundamental limitation of existing AI triage systems is their requirement for complete clinical data before generating predictions [23, 24]. `ETA` addresses this through two-phase operation generating risk estimates at distinct information availability stages.

#### 3.2.1 Phase 1: Pre-Triage Assessment

Phase 1 encompasses patient arrival through pre-vital prioritization under limited information.

##### Intake & Symptom Collection

The Intake Interview Agent (IIA) conducts adaptive conversational interviews at self-service kiosks, systematically collecting chief complaint, history of present illness (onset, location, duration, character, aggravating/alleviating factors, radiation, timing, severity), relevant medical history, medications, and allergies.

##### Pre-Vital Prioritization

The Clinical Reasoning Agent (CRA) analyzes symptom data to generate differential diagnoses and identify concerning patterns, retrieving relevant ESI handbook passages via retrieval-augmented generation (ChromaDB vector store with OpenAI embeddings). For complex presentations, the CRA queries PubMed API for literature validation. The Priority Assessment Agent (PAA) then synthesizes this analysis to generate: (i) tentative ESI estimate with confidence interval and (ii) queue priority score (HIGH/LOW). This enables clinicians to rank waiting patients by predicted urgency before vital sign collection.

#### 3.2.2 Phase 2: Post-Vital Classification

After clinicians collect vital signs and physical findings (appearance, skin condition, respiratory effort, mental status), the CRA performs comprehensive analysis integrating the complete clinical picture. The Triage Classification Agent (TCA) applies the ESI decision algorithm. Outputs include: final ESI recommendation, calibrated confidence score, structured rationale with explicit ESI criteria references, and uncertainty flags for cases requiring additional scrutiny.

### 3.3 Agent Specifications

#### Orchestrator Agent (OA)

Coordinates workflow execution, maintains session state, routes data between agents, and synthesizes outputs for dashboard presentation. Implements urgency bypass routing when IIA detects high-acuity indicators. Uses GPT-4.1-mini.

#### Intake Interview Agent (IIA)

Conducts patient-facing conversational symptom collection following structured clinical interview framework (OLDCARTS). Detects emergency indicators and triggers truncation when appropriate. Outputs structured JSON with chief complaint, HPI elements, history, medications, allergies, and emergency flags. Uses GPT-4.1-nano for reduced patient-facing latency.

#### Clinical Reasoning Agent (CRA)

Generates differential diagnoses, retrieves ESI guideline passages via RAG, and optionally queries PubMed for complex cases. Identifies red flags and risk factors. Operates in both phases with different input completeness. Uses GPT-4.1-mini with access to ChromaDB (ESI handbook) and PubMed API.

#### Priority Assessment Agent (PAA)

Generates pre-vital urgency estimates from symptoms alone. Produces tentative ESI levels with wide confidence intervals reflecting uncertainty, queue priority scores, and immediate assessment flags. Explicitly designed to operate conservatively under partial information. Uses GPT-4.1-mini.

#### Triage Classification Agent (TCA)

Produces final ESI recommendations with calibrated confidence and structured rationale explicitly referencing ESI criteria. Applies complete ESI decision algorithm after vital signs available. Uses GPT-4.1-mini.

#### Confidence Calibration

The TCA produces calibrated confidence estimates through a weighted ensemble of complementary evidence sources. The final confidence score, *C*_final_, is computed as:

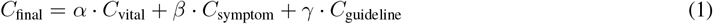

where *C*_vital_ represents confidence derived from physiologic stability based on vital signs, *C*_symptom_ captures confidence associated with symptom severity assessment, and *C*_guideline_ reflects alignment with Emergency Severity Index (ESI) handbook criteria. The weighting parameters *α, β, γ* satisfy *α* + *β* + *γ* = 1.

Initial weights (*α* = 0.5, *β* = 0.3, *γ* = 0.2) were selected as heuristic priors reflecting established clinical practice, in which physiologic instability is prioritized over symptom report alone, while explicit guideline alignment provides secondary confirmation. These weights are not assumed to be optimal; rather, they serve as a conservative initialization emphasizing patient safety. To assess robustness, we conducted a sensitivity analysis across multiple clinically plausible weight configurations (Appendix A.1), demonstrating that overall performance and safety characteristics remain stable, with no increase in undertriage.

For cases where confidence falls below threshold *τ* = 0.7, the system applies a conservative adjustment to reduce undertriage risk:

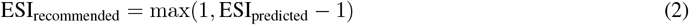

The threshold *τ* is treated as a policy parameter controlling the trade-off between specificity and clinical safety. We evaluated *τ* ∈ {0.6, 0.7, 0.8} and selected *τ* = 0.7 as the lowest threshold achieving zero undertriage across all evaluated cases while maintaining acceptable overall accuracy (Appendix A.2).

### 3.4 Technical Implementation

#### Prompt Engineering Strategies

All agent prompts employ: (i) explicit JSON output schemas with examples, (ii) clear role definitions and scope constraints, and (iii) clinical safety instructions (conservative uncertainty handling, red flag vigilance, deference to clinician judgment, hallucination prevention).

#### Knowledge Grounding

The RAG system processes the ESI Implementation Handbook v4 [4] into overlapping chunks, embeds using text-embedding-3-small, and stores in ChromaDB. At inference, the CRA generates queries based on chief complaint and symptoms, retrieving top-5 similar passages via cosine similarity for context integration. RAG enables updateability (modify corpus without retraining), transparency (display retrieved passages), and attribution (cite specific handbook sections).

### 3.5 Safety & Oversight Mechanisms

#### Preserved clinician Authority

`ETA` generates recommendations requiring active clinician acceptance; it never autonomously assigns ESI levels. Dashboard interface enables frictionless overrides with optional rationale capture. Override data supports both accountability and model improvement feedback.

#### Confidence Communication

TCA outputs calibrated confidence estimates with low-confidence classifications explicitly flagged. Structured rationale enables verification against clinical knowledge rather than opaque predictions.

#### Fail-Safe Design

System defaults conservatively: (i) low-confidence cases recommend higher acuity level, (ii) ESI-1/2 recommendations trigger automatic nursing alerts, (iii) component failures defer to manual triage with staff notification rather than unreliable recommendations.

#### Audit Logging

All operations logged to SQLite database capturing sessions, agent invocations, RAG retrievals, and decisions/overrides with timestamps, enabling complete reasoning chain reconstruction.

## 4 Evaluation

### 4.1 Evaluation Methodology

#### Dataset

The ESI Implementation Handbook v4 provides 60 standardized case studies comprising two distinct subsets [4]. The first 30 cases serve as practice cases designed for initial ESI training and familiarization with the decision algorithm. The second 30 cases function as competency assessment cases intended to evaluate clinician proficiency after training completion. This structure enables us to assess `ETA`’s performance on both routine training scenarios and more challenging competency validation cases that test nuanced clinical reasoning. Table 2 shows the distribution across ESI levels and case types. The dataset provides relatively uniform representation across acuity levels, enabling robust evaluation across the full triage spectrum.

**Table 2:** ESI Handbook evaluation dataset characteristics.

| ESI | #P | #C | Total | % | Chief Complaint | #P | #C | Total | % |
| --- | --- | --- | --- | --- | --- | --- | --- | --- | --- |
| ESI-1 | 6 | 6 | 12 | 20.00% | Chest Pain/Cardiac | 1 | 1 | 2 | 3.33% |
| ESI-2 | 8 | 6 | 14 | 23.33% | Abdominal Pain | 5 | 4 | 9 | 15.00% |
| ESI-3 | 6 | 7 | 13 | 21.67% | Respiratory | 1 | 2 | 3 | 5.00% |
| ESI-4 | 4 | 6 | 10 | 16.67% | Trauma/Injury | 6 | 8 | 14 | 23.33% |
| ESI-5 | 6 | 5 | 11 | 18.33% | Neurological | 5 | 4 | 9 | 15.00% |
|  |  |  |  |  | Other | 12 | 11 | 23 | 38.33% |

Note: #P = Practice cases (n=30); #C = Competency cases (n=30); Total N=60.

#### Simulation Protocol

Following AgentClinic methodology [29], we implemented automated evaluation using simulated agents providing standardized inputs. For each case: (i) a simulated patient agent (GPT-4.1-nano) responds to IIA queries by extracting symptom information from vignettes with naturalistic phrasing variation; (ii) after Phase 1, a simulated clinician agent (GPT-4.1-nano) extracts vital signs and physical findings for Phase 2; (iii) system outputs are compared against ground truth assignments. This approach ensures reproducible evaluation while introducing realistic linguistic variation.

#### Metrics

We evaluated performance using standard classification metrics adapted for triage. For Phase 1 (pre-vital assessment), we measured tentative ESI accuracy (exact match and within ±1 level) and binary priority classification performance (High [ESI 1–3] vs. Low [ESI 4–5]) with sensitivity and specificity. For Phase 2 (post-vital classification), we assessed final ESI accuracy, per-class performance, high-acuity (ESI 1–2) detection rates, and clinical safety through undertriage and overtriage rates (defined as misclassifications ≤ 2 levels). We also measured processing latency (mean, median, 95th percentile) for both phases and end-to-end operation.

#### Statistical Uncertainty Estimation

Given the limited sample size (n = 60), all performance metrics were estimated using non-parametric bootstrap resampling at the case level. We generated 1000 bootstrap samples by sampling with replacement from the original case set and recomputed all evaluation metrics for each sample. We report point estimates along with 95% confidence intervals.

### 4.2 Results

#### Phase 1: Pre-Vital Prioritization Performance

Table 3 presents Phase 1 performance operating on symptom data alone, without vital signs or physical examination findings. Despite operating under substantial information constraint, `ETA` achieved 66.67% (95% CI: 55.00%-78.33%) exact ESI match and 95.00% (95% CI: 88.33%-100.00%) within ±1 accuracy. The 28.33% gap between exact and near-exact accuracy reflects inherent uncertainty when vital signs are unavailable, a fundamental limitation acknowledged in ESI guidelines, which prohibit definitive classification without physiologic data.

**Table 3:** Phase 1 pre-vital performance.

| Metric | Practice | Competency | Combined |
| --- | --- | --- | --- |
| *Tentative ESI Classification* |  |  |  |
| Exact Match Accuracy | 66.67% [49.92%, 83.33%] | 66.67% [49.92%, 83.33%] | 66.67% [55.00%, 78.33%] |
| Within-$\pm 1$ Accuracy | 93.33% [83.33%, 100.00%] | 96.67% [90.00%, 100.00%] | 95.00% [88.33%, 100.00%] |
| *Priority Classification* |  |  |  |
| Overall Accuracy | 90.00% [80.00%, 100.00%] | 83.33% [70.00%, 93.42%] | 86.67% [76.67%, 95.00%] |
| High Priority Sensitivity | 90.00% [73.68%, 100.00%] | 84.21% [73.33%, 100.00%] | 87.18% [76.86%, 97.44%] |
| High Priority Specificity | 90.00% [72.73%, 100.00%] | 81.82% [67.65%, 100.00%] | 85.71% [66.67%, 100.00%] |

More critically, for the binary prioritization task determining which patients require immediate vital sign assessment, the system achieved 86.67% (95% CI: 76.67%-95.00%) accuracy with 87.18% (95% CI: 76.86%-97.44%) sensitivity for high-priority cases (ESI 1–3). This sensitivity indicates the system correctly identified 34 of 39 patients requiring urgent evaluation, with only 5 high-acuity patients initially misclassified as low priority. The 85.71% (95% CI: 66.67%-100.00%) specificity demonstrates appropriate restraint, avoiding excessive false alarms that would undermine clinician trust.

#### Phase 2: Final Classification Performance

Table 4 presents Phase 2 performance with complete clinical information including vital signs and physical findings. With complete data, `ETA` achieved 80.00% (95% CI: 70.00%-92.35%) exact match accuracy and 98.33% (95% CI: 95.33%-100.00%) within ±1 accuracy, a 13.33 percentage point improvement over Phase 1 exact match, demonstrating effective integration of vital sign data. High-acuity sensitivity of 92.31% (95% CI: 80.77%-100.00%) (24/26 ESI 1–2 cases correctly identified) indicates reliable detection of patients requiring immediate intervention. Only two ESI-2 cases were misclassified as ESI-3.

**Table 4:** Phase 2 post-vital performance.

| Metric | Practice | Competency | Combined |
| --- | --- | --- | --- |
| *ESI Classification Accuracy* |  |  |  |
| Exact Match | 86.67% [73.33%, 96.67%] | 73.33% [56.67%, 86.67%] | 80.00% [70.00%, 92.35%] |
| Within-±1 Level | 96.67% [88.73%, 100.00%] | 100.00% [100.00%, 100.00%] | 98.33% [95.33%, 100.00%] |
| *High-Acuity Detection* |  |  |  |
| Sensitivity | 100.00% [100.00%, 100.00%] | 83.33% [58.33%, 100.00%] | 92.31% [80.77%, 100.00%] |
| Specificity | 81.25% [62.50%, 93.91%] | 83.33% [66.67%, 100.00%] | 82.35% [67.65%, 94.12%] |
| *Clinical Safety* |  |  |  |
| Undertriage Rate | 0.00% [0.00%, 0.00%] | 0.00% [0.00%, 0.00%] | 0.00% [0.00%, 0.00%] |
| Overtriage Rate | 3.33% [0.00%, 6.67%] | 0.00% [0.00%, 0.00%] | 1.67% [0.00%, 3.33%] |

Performance stratification revealed higher accuracy on practice cases (86.67% (95% CI: 73.33%-96.67%)) compared to competency cases (73.33% (95% CI: 56.67%-86.67%)), as expected given the latter’s design for proficiency assessment with more complex presentations. Notably, practice cases achieved perfect high-acuity sensitivity (100.00% (95% CI: 100.00%-100.00%)) while competency cases demonstrated 83.33% (95% CI: 58.33%-100.00%), though both maintained the critical 0.00% (95% CI: 0.00%-0.00%) undertriage rate.

Competency cases achieved perfect within-±1 accuracy (100.00% (95% CI: 100.00%-100.00%)), indicating that while exact ESI assignment was more challenging for complex presentations, the system consistently remained within one acuity level of ground truth.

From a clinical safety perspective, the system demonstrated excellent safety profile with 0.00% (95% CI: 0.00%-0.00%) undertriage rate (no cases misclassified ≥ 2 levels below ground truth) and minimal 1.67% (95% CI: 0.00%-3.33%) overtriage rate (1/60 cases ≥2 levels above). This near-zero error rate for clinically dangerous misclassifications, combined with conservative bias favoring overtriage, aligns with clinical practice where the cost of missing critical cases far exceeds unnecessary resource utilization.

Table 5 presents per-class accuracy. ESI-1 achieved perfect classification (100.00% (95% CI: 100.00%-100.00%)), while ESI-2 demonstrated 85.71% (95% CI: 64.29%-100.00%) accuracy. ESI-3 performance (61.54% (95% CI: 38.46%-84.62%)) reflects well-documented inter-rater variability at ESI-2/3 and ESI-3/4 boundaries where clinical presentations overlap and resource prediction becomes subjective. ESI-4 achieved robust 90.00% accuracy. ESI-5 demonstrated lower accuracy (63.64% (95% CI: 36.36%-90.91%)), with 4/11 cases over-triaged to ESI-4, likely reflecting conservative system bias toward predicting resource utilization in ambiguous non-urgent presentations. Misclassifications predominantly occurred between adjacent levels rather than across multiple categories.

**Table 5:**
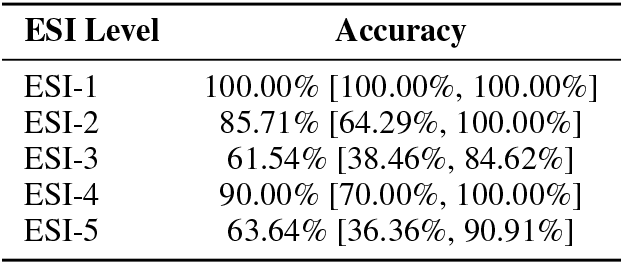
Per-class classification accuracy by ESI level.

#### System Latency

Table 6 presents processing times. Mean end-to-end latency of 48.95 seconds comprises 36.14 seconds for Phase 1 (pre-vital assessment) and 13.70 seconds for Phase 2 (post-vital classification). The 95th percentile latencies (47.17 sec Phase 1, 17.32 sec Phase 2, 60.25 sec end-to-end) indicate worst-case performance remains within clinical acceptability. Current ED triage typically requires 2–5 minutes per patient; `ETA`’s latency profile enables integration without workflow disruption, particularly if Phase 1 intake occurs concurrently with patient arrival processing.

**Table 6:** Processing latency across evaluation runs.

| Metric | Mean | Median | 95th %ile |
| --- | --- | --- | --- |
| Phase 1 Latency | 36.14 sec | 36.88 sec | 47.17 sec |
| Phase 2 Latency | 13.70 sec | 13.17 sec | 17.32 sec |
| End-to-End | 48.95 sec | 49.08 sec | 60.25 sec |

### 4.3 Error Analysis

We analyzed all 12 Phase 2 misclassifications (20% error rate across 60 cases) to identify systematic failure modes and their underlying mechanisms.

#### Resource prediction errors

The dominant failure mode involved ESI-3/4/5 boundary confusion, where the system incorrectly anticipated the number of diagnostic resources required for stable patients. For instance, a lip laceration requiring conscious sedation (ground truth: ESI-3, two resources) was classified as ESI-4, under-estimating procedural complexity by counting only the repair without accounting for sedation and monitoring. These errors reflect inherent ambiguity in prospective resource forecasting, a limitation acknowledged in ESI training materials for human triage nurses, particularly when distinguishing between single procedures versus procedures requiring ancillary resources.

#### Vital sign over-interpretation

The second major failure mode involved over-triaging stable patients based on single abnormal vital signs without adequate clinical contextualization. The system’s deterministic vital sign assessment flags borderline abnormalities as “HIGH-RISK” without accounting for physiologic context. For example, a toothache case with pain severity 10/10 but otherwise normal vitals was classified as ESI-2, with severe pain misinterpreted as “severe distress” warranting emergent care. An open fracture with normal vital signs (ground truth: ESI-3) was over-triaged to ESI-2 based on injury severity alone. While this conservative bias contributes to high-acuity sensitivity and zero undertriage rate, it reduces classification precision for stable patients with isolated abnormal findings.

#### High-risk presentation under-recognition

Most concerning were two clinically significant misclassification cases where high-risk ESI-2 scenarios were misclassified as ESI-3, despite meeting ESI Handbook criteria for emergent care. A syncopal episode in a 16-year-old with normal vital signs was classified as ESI-3, with the system failing to recognize syncope as a high-risk presentation requiring comprehensive workup regardless of current stability. Similarly, chest pain in a young adult with normal vitals was undertriaged to ESI-3, reflecting under-weighting of cardiac risk factors when vital signs are reassuring. These cases highlight a critical limitation: the system’s difficulty in identifying high-risk presentations based on symptomatology alone when vital signs provide false reassurance. While rare, such errors represent safety-critical failures where patients requiring immediate evaluation were assigned lower priority.

#### Phase 1 specific patterns

Analysis of Phase 1 confusion patterns revealed systematic over-triage in the absence of vital signs. ESI-2 and ESI-3 cases were frequently over-triaged to higher acuity levels, reflecting conservative estimation when lacking physiologic data. This pattern serves the clinical goal of ensuring high-risk patients receive expedited vital sign assessment during queue management, even at the cost of lower specificity. Phase 2 accuracy improved substantially with vital sign availability (80.00% vs 66.67%), demonstrating that physiologic data enables more precise classification while maintaining safety (0.00% undertriage rate).

These error patterns suggest refinement opportunities: (1) enhanced logic for counting procedural resources with explicit sedation/monitoring consideration, (2) context-aware vital sign interpretation incorporating age-specific norms and clinical stability indicators, and (3) strengthened symptom-based risk stratification to identify high-risk presentations independent of vital sign reassurance.

### 4.4 Evaluation Limitations

Our simulation based evaluation relies on standardized ESI handbook vignettes and simulated patient and clinician agents, enabling reproducible benchmarking but limiting ecological validity. This setup does not fully capture real-world emergency department conditions, including variability in patient communication, health literacy, and language proficiency; communication barriers such as accents, speech impairments, or emotional distress; system integration challenges. Additionally, ESI handbook cases may not reflect true ED presentation distributions or the degree of ambiguity encountered in practice, and ground truth ESI labels represent single authoritative classifications where reasonable experts may disagree. Consequently, system performance in live clinical environments, where information may be incomplete, delayed, or inconsistently communicated, may differ from observed simulation results, motivating prospective clinical evaluation.

## 5 Discussion

We present `ETA`, a multi-agent AI framework addressing a critical gap in emergency triage decision support, the inability of existing systems to support prioritization decisions under partial information. Our two-phase architecture generates actionable risk estimates both pre-vital (for queue management) and post-vital (for final classification), while preserving clinician decision authority through interpretable, criterion linked reasoning.

### 5.1 Key Contributions

#### Two-Phase Workflow for Sequential Decision Support

Unlike prior systems requiring complete clinical data [23, 24], `ETA` mirrors triage’s inherently sequential nature by generating calibrated risk estimates at distinct information availability stages. Phase 1 pre-vital assessment enables clinicians to identify which waiting patients require immediate evaluation during high-volume periods, a capability absent in existing approaches. Our evaluation demonstrates 87.18% (95% CI: 76.86%-97.44%) high-priority sensitivity, suggesting the system can reliably flag patients requiring expedited assessment from symptom data alone.

#### Urgency Adaptive Interview Strategy

The Intake Interview Agent implements dynamic question truncation when high-acuity indicators are detected, balancing comprehensive data collection against the imperative to rapidly identify life threatening presentations. This design acknowledges that optimal triage support varies with presentation urgency, a nuance missing from fixed-depth interview protocols.

#### Interpretable Multi-Agent Architecture

By decomposing triage into specialized, collaborative agents with explicit reasoning chains linked to ESI criteria, `ETA` enables verification of system logic against professional knowledge. This transparency supports appropriate trust calibration and facilitates identification of cases where system reasoning may be incomplete, addressing known adoption barriers for clinical AI [13, 25].

### 5.2 Deployment Considerations

Successful clinical integration of AI systems requires comprehensive planning that extends beyond technical development to address organizational readiness, stakeholder engagement, and ethical governance [30]. Clinical translation requires addressing practical constraints beyond algorithmic performance:

#### Integration Challenges

EHR interoperability via FHIR APIs, workflow analysis to ensure augmentation rather than disruption, and staff training on appropriate reliance calibration are prerequisites. Cost-benefit analysis should weigh API expenses (estimated $0.76 per 100 encounters based on 20,563 tokens/case) against potential benefits such as reduced triage time, improved consistency, and enhanced safety.

#### Speech-Enabled and Multilingual Deployment

Real-world deployment should incorporate speech-to-text (STT) and text-to-speech (TTS) interfaces and multilingual LLM support to enable accessible, language-agnostic symptom intake in diverse emergency departments. Prior work has demonstrated that foundation models can be applied directly in non-English languages without additional pretraining or fine-tuning [12, 31]. The effects of these capabilities on accuracy, latency, and equity require prospective evaluation.

#### Safety and Oversight

`ETA` preserves clinician decision authority with all outputs framed as recommendations requiring active review. Conservative uncertainty handling defaults to higher acuity when confidence is low. Comprehensive audit logging supports accountability and retrospective review. Fail-safe design ensures graceful degradation to manual triage during component failures.

#### Equity Monitoring

Our ESI Handbook based evaluation lacks demographic stratification, leaving potential performance disparities across patient populations unexamined. Clinical deployment should include disaggregated performance monitoring, threshold-based disparity alerts, and periodic equity audits with stakeholder input to identify and remediate biases.

### 5.3 Limitations and Future Work

This study has several limitations that motivate future investigation. First, evaluation using simulated patients and standardized cases may not fully capture the complexity of real clinical communication, including incomplete or contradictory symptom descriptions, variations in health literacy, and the dynamic nature of emergency department interactions; consequently, performance in real-world deployment may differ from simulation results. Second, evaluation on the ESI Handbooks dataset may not reflect true emergency department presentation distributions and may overrepresent relatively clear cases compared to the ambiguity encountered in clinical practice, highlighting the need for multi-site evaluation across diverse patient populations.

Finally, while this work demonstrates technical feasibility, it does not establish clinical utility. Translational challenges in real-world deployment warrant careful consideration, including whether the system addresses genuine clinical needs, integrates seamlessly into existing workflows, and maintains performance across diverse patient populations [14]. Prospective evaluation in emergency department settings is therefore required to assess impact on triage accuracy and consistency relative to standard practice, effects on clinician cognitive load and workflow efficiency, performance across demographic groups to identify potential disparities, and clinician acceptance with appropriate trust calibration.

## 6 Conclusion

Emergency department triage operates under time pressure, information uncertainty, and high stakes, where rapid acuity assessment directly impacts patient outcomes. Rising volumes, increasing complexity, and persistent staffing challenges motivate AI-assisted approaches that augment clinical decision making while preserving professional nursing judgment.

We presented `ETA`, a multi-agent framework addressing a critical gap in triage decision support, existing systems require complete clinical data before operation, failing to support the sequential, information gathering reality where patients accumulate before vital signs are collected. `ETA`’s two-phase architecture generates actionable risk estimates both pre-vital (enabling queue prioritization from symptoms alone) and post-vital (providing final ESI recommendations with complete information), while preserving clinician authority through interpretable, criterion linked reasoning.

Evaluation on 60 standardized triage cases demonstrates 86.67% (95% CI: 76.67%-95.00%) priority classification accuracy in Phase 1 with 87.18% (95% CI: 76.86%-97.44%) high-priority sensitivity, and 80.00% (95% CI: 70.00%-92.35%) final ESI classification accuracy in Phase 2. The multi-agent architecture, comprising specialized agents for intake, clinical reasoning, prioritization, and classification, provides modularity, transparency, and interpretability advantages over monolithic approaches. RAG grounds recommendations in ESI handbook criteria, enabling explicit guideline citation and clinician verification.

This work demonstrates feasibility of multi-agent AI for time-critical clinical decisions under partial information. Prospective evaluation in emergency departments is essential to validate clinical utility, assess real-world performance across diverse patient populations. The two-phase design principle, generating useful predictions from incomplete information while refining as data accrues, may inform decision support for trauma evaluation, deterioration monitoring, and telemedicine screening where timely decisions cannot await complete data collection.

## Data Availability

All data produced in the present work are contained in the manuscript

https://github.com/Karthick47v2/ED-Triage-Agent.

## A Sensitivity Analysis of Confidence Calibration Parameters

We evaluated the robustness of `ETA`’s performance to variations in confidence calibration parameters through systematic perturbation analysis. Weight configurations were selected to reflect clinically plausible emphasis on physiologic stability (*α*), symptom severity (*β*), and guideline alignment (*γ*), subject to the constraint *α* + *β* + *γ* = 1.

### A.1 Weight Configuration Sensitivity

Table 7 presents Phase 2 performance across five weight configurations.

**Table 7:**
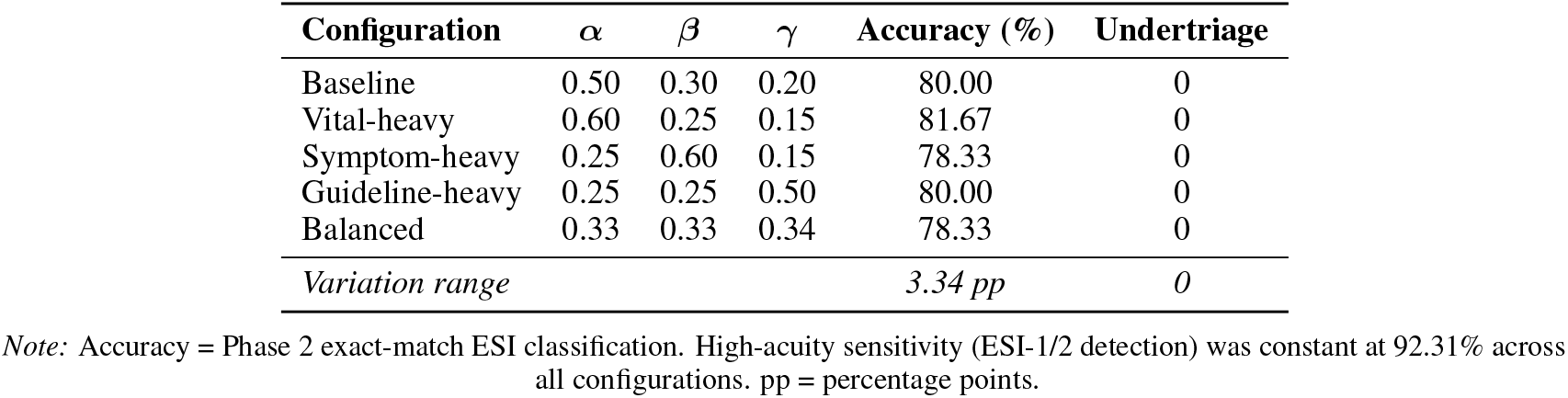
Performance robustness across confidence weight configurations.

Five configurations spanning the clinically relevant parameter space were evaluated on the full 60-case dataset (Table 7). Exact-match accuracy varied by only 3.34 percentage points, while high-acuity sensitivity remained invariant at 92.31% across all configurations. Critically, zero undertriage events occurred under any weight configuration, demonstrating robust safety characteristics independent of parameter selection.

### A.2 Confidence Threshold Sensitivity

Table 8 presents performance across three candidate thresholds. The threshold *τ* controls application of the conservative adjustment (Equation 2), creating a fundamental trade-off between classification accuracy and clinical safety.

**Table 8:** Accuracy-safety trade-off across confidence threshold values.

| Threshold ($\tau$) | Accuracy (%) | Undertriage | Overtriage |
| --- | --- | --- | --- |
| 0.60 | 83.33 | 1 | 0 |
| 0.70 | 80.00 | 0 | 1 |
| 0.80 | 71.67 | 0 | 3 |

Note: Undertriage and overtriage denote misclassifications $\geq 2$ ESI levels. Within- $\pm 1$ accuracy remained constant at 98.33% across all thresholds. High-acuity sensitivity was invariant at 92.31%.

Threshold *τ* = 0.60 maximized accuracy (83.33%) but introduced a critical undertriage event (ESI-2 to ESI-4), violating safety requirements. Conversely, *τ* = 0.80 eliminated undertriage but incurred excessive overcalling (three overtriage events). We selected *τ* = 0.70 as the optimal operating point, representing the lowest threshold achieving zero undertriage while maintaining acceptable accuracy (80.00%) with minimal overcalling (1.67%).

These results demonstrate that `ETA`’s performance characteristics remain stable across reasonable parameter variations, with safety (zero undertriage) maintained throughout the clinically plausible parameter space. The conservative design philosophy achieves its primary objective, preventing dangerous undertriage without requiring precise parameter tuning.

